# Validation of the VACTERL Episignature and Evidence for Epigenomic Convergence Across Recurrent Constellations of Embryonic Malformations

**DOI:** 10.64898/2026.07.10.26357391

**Authors:** Julianne Postma, Sadegheh Haghshenas, Teija M. I. Bily, Majdina Isovic, Alexandre White-Brown, Haley McConkey, Jennifer Kerkhof, Jessica Rzasa, Maha Saleh, Chitra Prasad, Victoria M. Siu, Melissa T. Carter, David A. Dyment, Joanna Lazier, Sarah L. Sawyer, Angelica A. Moresco, Maria Jimena Diaz, Silvina L. Abbate, Philippe M. Campeau, A. Micheil Innes, Kym M. Boycott, Bekim Sadikovic, Tugce B. Balci

## Abstract

**Background:** Recurrent constellations of embryonic malformations (RCEMs) comprise multiple malformation conditions with largely unexplained etiologies and no established molecular biomarkers. A shared DNA methylation episignature was recently identified in VACTERL association and oculoauriculovertebral spectrum (OAVS). We sought to validate this episignature in an independent, deeply phenotyped cohort and evaluate its detection across related RCEMs.

**Methods:** Genome-wide DNA methylation profiling was performed on peripheral blood from 38 participants with clinically diagnosed RCEMs, including VACTERL (n=21), partial VACTERL (n=3), OAVS (n=3), and other RCEM-related conditions (n=11).

**Results:** The Episign V5 RCEM episignature demonstrated robust sensitivity for VACTERL (18/21, 85.7% positive), while the remaining three participants showed intermediate positivity. Of three participants with partial VACTERL, one with tracheoesophageal fistula demonstrated intermediate positivity, whereas the other two were negative. Episignature positivity was also identified in oculoauriculofrontonasal dysplasia (1/1, robust) and rhomboencephalosynapsis (1/2, robust) but was limited in OAVS (1/3, intermediate) and absent in frontonasal dysplasia (0/4).

**Conclusions:** Independent validation establishes the Episign V5 RCEM episignature as a reproducible molecular biomarker for VACTERL, a condition that remains a diagnosis of exclusion. Variable detection across related malformation conditions suggests etiologic heterogeneity, whereas overlap among selected phenotypes supports epigenomic convergence across the RCEM spectrum.

## INTRODUCTION

The term “recurrent constellations of embryonic malformations” (RCEM) was first proposed by Adam et al. (2020) to describe a group of multiple malformation associations or syndromes presumed to share an early embryonic origin for which no genetic cause has been identified. Disorders initially proposed as RCEMs include limb body wall complex (LBWC), Müllerian duct aplasia-renal anomalies-cervicothoracic somite dysplasia (MURCS), oculoauriculovertebral spectrum (OAVS), omphalocele-exstrophy-imperforate anus-spinal defects (OEIS) complex, pentalogy of Cantrell (POC), and vertebral-anal-cardiac-tracheoesophageal fistula-renal-limb (VACTERL) association (Adam et al., 2020). These conditions share several characteristic clinical features, including relative commonality (reported in >150 individuals), minimal familial recurrence, a statistically significant association with monozygotic twinning (with approximately 80% discordance for phenotype), negative genetic testing, and typically preserved intellectual function in survivors (Adam et al., 2020). Other unsolved conditions, such as PHACES (Posterior Fossa, Brain malformations, hemangiomas, arterial anomalies, cardiac defects, eye anomalies, sternal clefting/supraumbilical raphe) association and rhomboencephalosynapsis (RES) were not initially included due to a female preponderance and the association of intellectual disability, respectively (Adam et al., 2020). However, these and other conditions including sirenomelia and urorectal septum malformation sequence (URSMS) have been proposed to fall within the RCEM spectrum (Stevenson et al., 2021, Mark et al., 2022).

Pathologic mechanisms underlying both defined and proposed RCEMs have been debated in the literature. Stevenson (2021) proposed “vitelline vascular steal” as a unifying mechanism for sirenomelia, OEIS complex, LBWC, and other caudal malformations, while Mark (2022) suggested NAD⁺ deficiency based on phenotypic overlap with genetic NAD⁺ biosynthesis disorders caused by biallelic variants in *KYNU*, *HAAO*, and *NADSYN1*. Disrupted methylation itself is associated with factors co-occurring with RCEMs, including monozygotic twinning (van Dongen et al., 2021) and assisted reproductive technology (ART), with an 18.5% rate of ART use reported among affected twin pregnancies (Adam et al., 2020). Collectively, these theories support the concept that RCEMs may represent a spectrum of malformations with shared epigenomic disruption, which reflects shared perturbation of core developmental programs.

A significant breakthrough in understanding RCEM pathogenesis was the description of an episignature as a biomarker for VACTERL and OAVS through DNA methylation analysis (EpiSign) (Haghshenas et al., 2024). A common episignature, with robust and intermediate subgroups, was identified spanning both VACTERL and OAVS phenotypes, as well as cases with overlapping features (Haghshenas et al., 2024). The robust and intermediate subgroups were defined based on their proximity to the control samples, where the robust group exhibited a more pronounced difference in methylation patterns from the controls.

DNA methylation arrays have rapidly emerged as powerful diagnostic tools in rare genetic conditions for detecting unique genome-wide patterns of aberrant methylation known as episignatures (Levy et al., 2021, Kerkhof et al., 2024). Episignatures have proven to be stable and reliable biomarkers in clinical settings, facilitating the diagnosis of rare genetic disorders, reclassification of variants of uncertain significance, and molecular characterization of clinically heterogeneous conditions (Kerkhof et al., 2024). They have now been defined for more than 120 genetic disorders and were recently expanded to also have application in non-genetic and environmental conditions, such as fetal valproate syndrome and fetal alcohol spectrum disorder (Levy et al., 2021, Van der Laan et al., 2025, Haghshenas et al., 2024). This has broadened the conceptual scope of episignatures beyond their origins in Mendelian disorders of the epigenetic machinery, establishing them as stable molecular records of functional disruption during early development; whether caused by variants in genes within or outside the classical epigenetic machinery or, in some cases, by environmental exposures (Tkemladze et al, 2025).

We sought to validate the Episign V5 RCEM episignature in an independent VACTERL cohort, to refine its performance in OAVS, and to extend this analysis to an expanded RCEM-spectrum to investigate potential episignature overlap. By moving beyond gene-centric models, episignature profiling may establish reproducible biomarkers for conditions historically diagnosed by exclusion, define biologically coherent subgroups within clinically heterogeneous malformation spectra, and provide a framework for studying disruption of human development.

## METHODS

### Participant Recruitment and Characterization

This cohort included DNA from peripheral blood from 38 participants with a clinical diagnosis of an RCEM who were recruited from clinicians through existing multicentre national and international collaborations from several institutions: London Health Sciences Centre (LHSC, London Ontario, Canada) (n=20), the Children’s Hospital of Eastern Ontario (CHEO, Ottawa, Ontario Canada) (n=10), Centre Hospitalier Universitaire Sainte-Justine (Montreal, Quebec, Canada) (n=1) and National Pediatric Hospital J. P. Garrahan (Buenos Aires, Argentina) (n=7). Two of the patients enrolled through Care4Rare Canada were from international centres, including Hunter New England Health (Newcastle New South Wales, Australia), and Hospital de Reabilitação de Anomalies Craniofaciais (Sao Paulo, Brazil). Two additional participants with multiple congenital anomalies, initially thought to represent an RCEM subtype, then diagnosed with monogenic conditions through sequencing (n=2), were also recruited to compare their episignatures with that of RCEMs (Table 1).

**Table 1.**
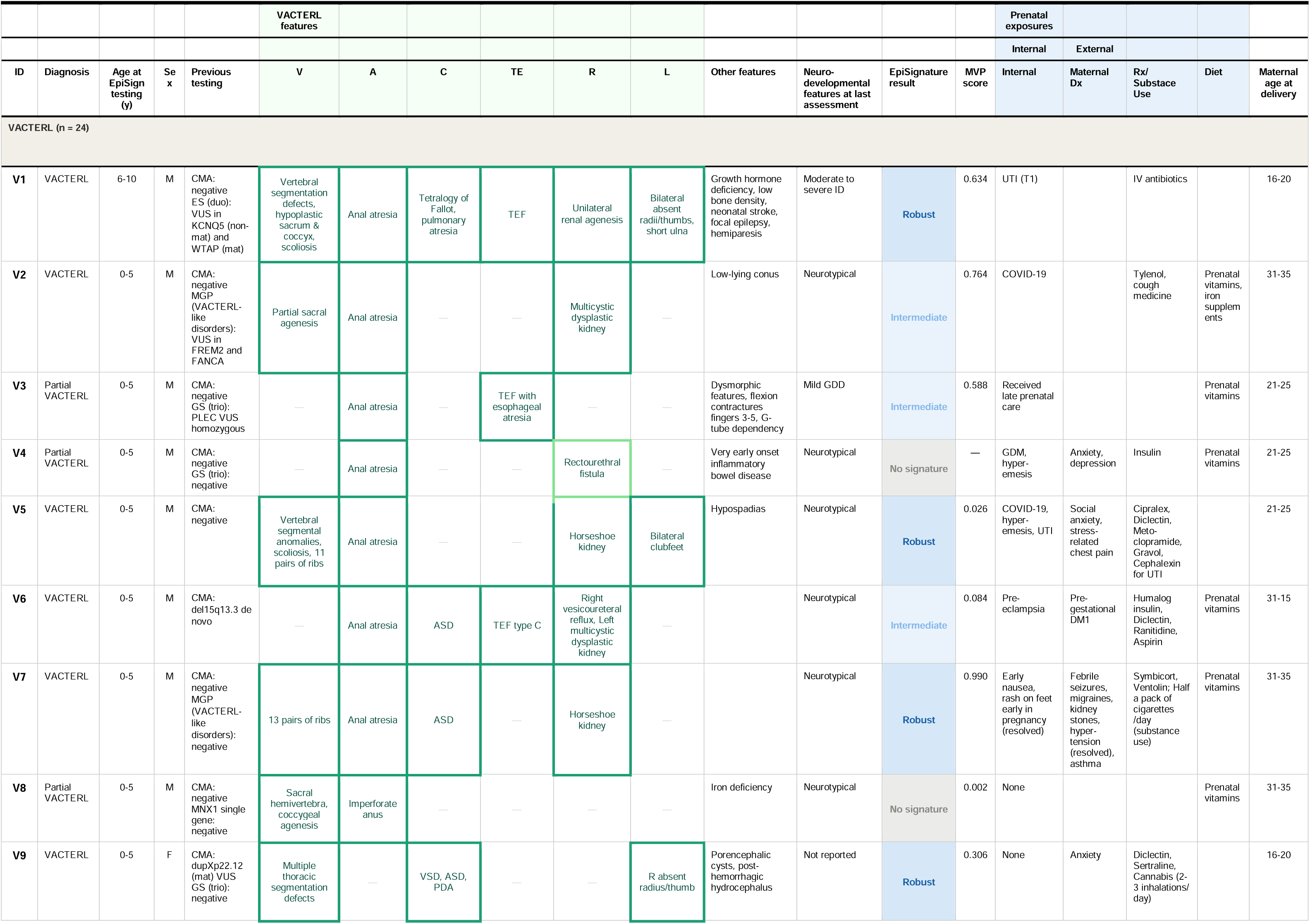
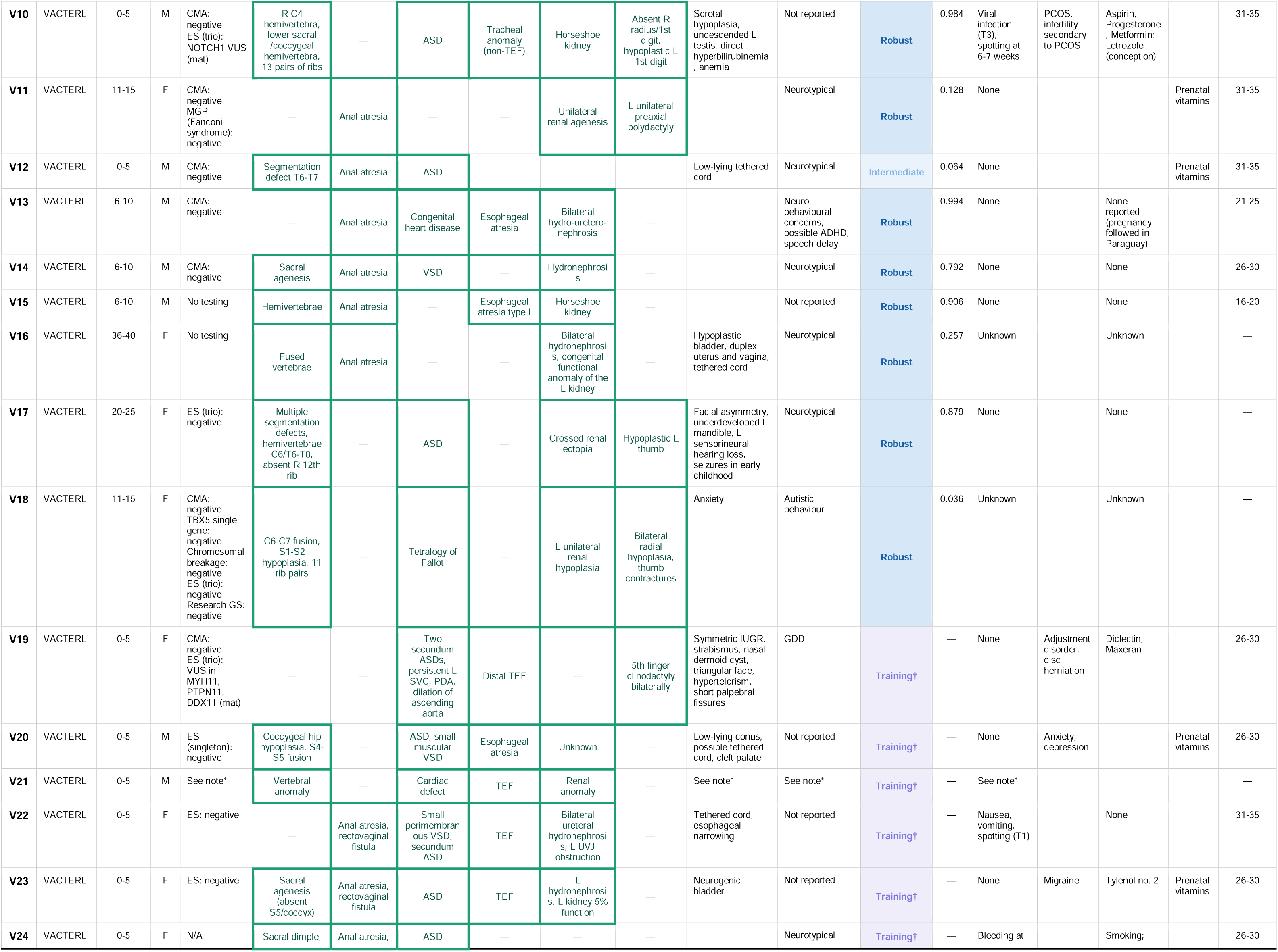
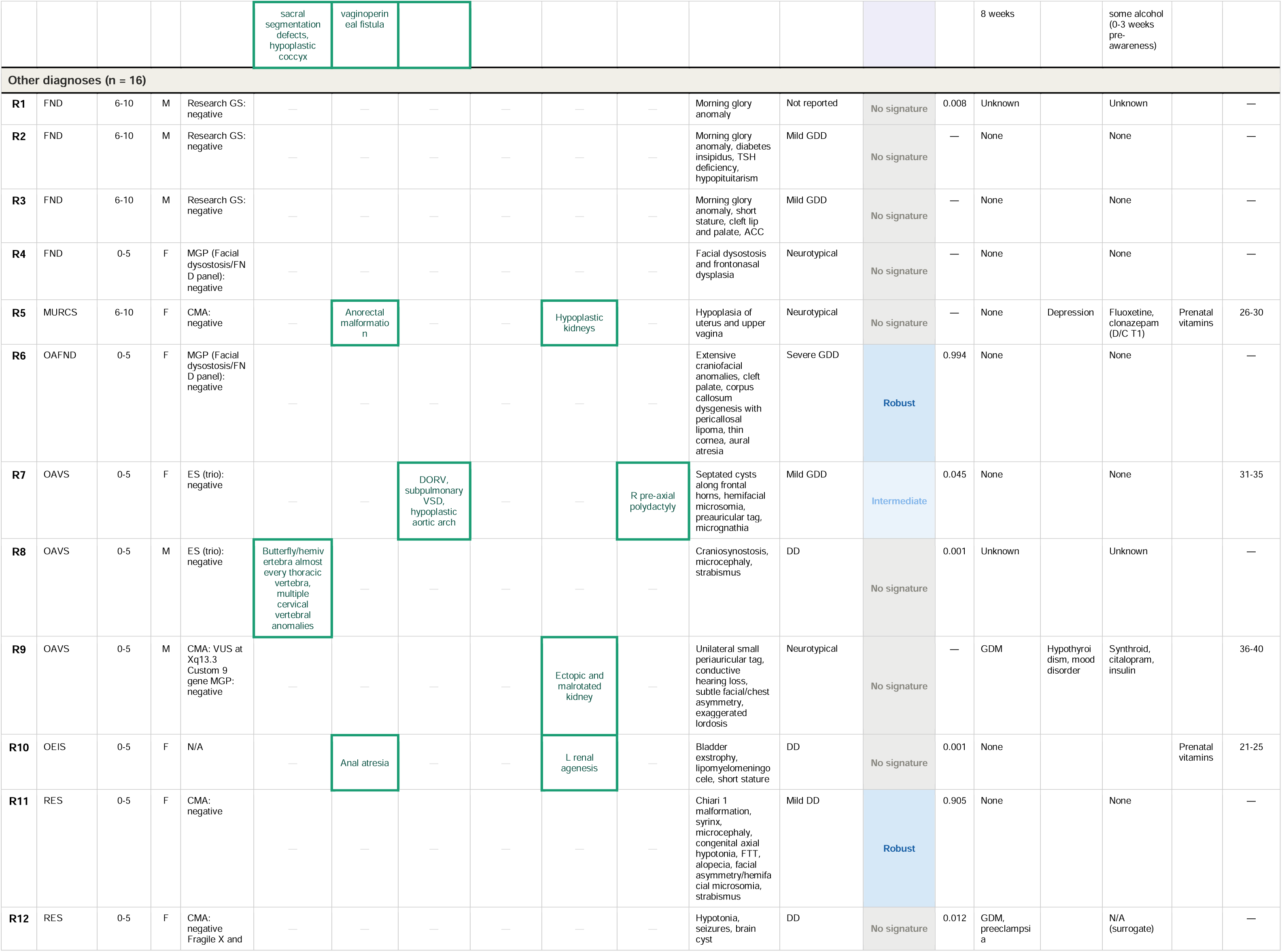
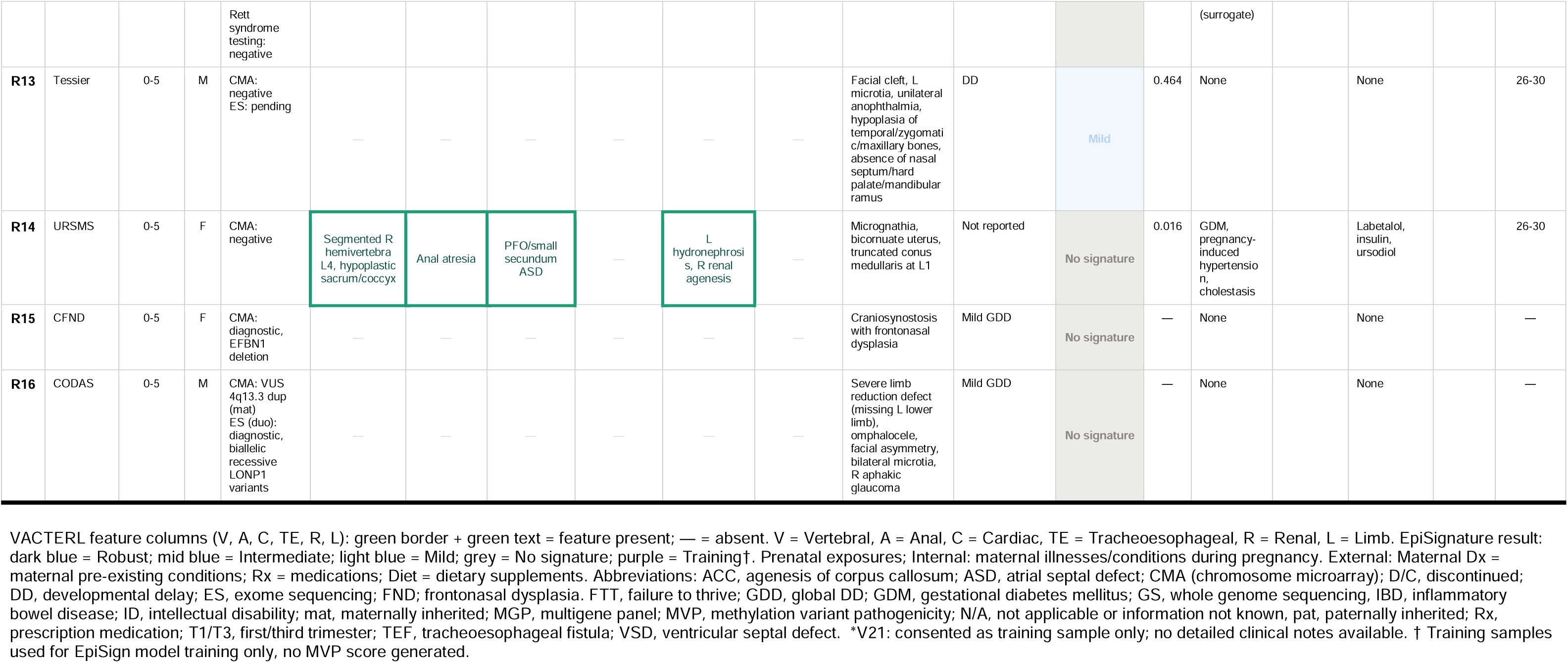
Patient cohort characteristics.

Informed consent for gathering of clinical and genomic data, as well as for the episignature studies was obtained from all participants or their guardians through a standardized research consent protocol approved by either the Care4Rare Canada consortium (CHEO Research Ethics Board:11/04E and CTO-1577), or the Episignature Discovery Study (Western University Health Sciences Research Ethics Board: HSREB #106302), or approved studies at local institutions’ research ethics boards. All samples were de-identified prior to testing. All participants were deeply phenotyped by clinical geneticists (MS, CP, VMS, AM, MTC, DAD, JL, SLS, MJD, PMC, AMI, KMB, TBB).

Regarding the clinical diagnostic process for an RCEM, a diagnosis of VACTERL was defined as the presence of at least 3 of 6 classical components; three individuals with only 2 features and thus classified as partial VACTERL (van de Putte et al., 2019) were also included for exploration. For other RCEM, published diagnostic criteria were used if present, and if not, participants were classified based on clinical judgement of overlap with published phenotypic features, such as those that are included in Adam et al., 2020.

Prenatal exposure data were gathered from medical records where available. Exposures were classified as “internal”, “specific external”, or “general external” as defined in the work from Wild, 2012, in which the exposome is subdivided into these groups based upon the nature of the exposure. Internal is defined as conditions or processes intrinsic to the body, such as metabolism and body morphology, while specific external involves exposures such as tobacco, radiation, medications, and other contaminants. General external is the broader environment of a person, such as social capital and education (Wild et al., 2012).

### DNA Methylation Analysis

In this work, we validated and tested the EpiSign V5 RCEM episignature and classifier, with original episignature development described in Haghshenas et al., 2024. Briefly, the EpiSign V5 episignature was identified using DNA from peripheral blood from participants with a clinical diagnosis of VACTERL or OAVS. DNA methylation data was generated using the Illumina Infinium Methylation EPIC BeadChip platform (Illumina, San Diego, CA), which assesses approximately 860,000 CpG loci genome-wide. Selected probes were identified through comparison with age and sex matched controls to identify the EpiSign V5 RCEM episignature.

The cohort described herein was tested against the EpiSign V5 RCEM episignature. Six of the 24 participants with VACTERL or partial VACTERL demonstrated a strong concordance with the episignature and were therefore included in the EpiSign V5 episignature training set. The rest of the VACTERL cohort (18/24) were used as validation participants. The participants with the following diagnoses were used as testing: 4 frontonasal dysplasia (FND), 3 OAVS, 2 RES, 1 MURCS, 1 URSMS, 1 oculo-auriculo-frontonasal dysplasia (OAFND), 1 OEIS, 1 Tessier cleft, 1 craniofrontonasal dysplasia (CFND), and 1 CODAS (cerebral, ocular, dental, auricular, skeletal) syndrome. Using the episignature probes, hierarchical clustering and multidimensional scaling (MDS) plots were generated, and a support vector machine (SVM) classifier was used to calculate methylation variant pathogenicity (MVP) scores ranging from 0 to 1, indicating the probability of a sample matching the EpiSign v5 RCEM episignature. To determine whether a sample matched the episignature, all three analyses were considered: whether the sample clustered with participants in the hierarchical clustering heatmap, whether it clustered with participants in the MDS plot, and whether it received an elevated MVP score. For a more detailed explanation of the interpretation of the clustering plots and MVP scores, please see Kerkhof et al., 2024.

## RESULTS

### Cohort demographics and clinical features

The study cohort consisted of 40 participants; of which 38 had either a classic RCEM or a diagnosis from an “expanded RCEM” list consisting of similar conditions: 21 with VACTERL, three with partial VACTERL, four with frontonasal dysplasia (FND), three with OAVS, two with RES, and one each with MURCS, URSMS, OAFND, OEIS, Tessier cleft; and two had monogenic conditions: one with CFND, and one with CODAS syndrome (Table 1). Sex distribution included 22 males and 18 females. All participants were unrelated, except for one mother-child pair.

Of the participants with VACTERL or partial VACTERL, where information on neurodevelopmental level was available (n=14), three were reported to have neurodevelopmental diagnoses (21%), including ADHD (n=1), autistic behaviour (n=1), and global developmental delay (GDD)/Intellectual disability (ID) (n=1). Of the other participants with RCEM, neurodevelopmental delay was reported in OAFND (n=1), OAVS (n=1), RES (n=2), and Tessier cleft (n=1) (Table 1).

### Prenatal History

Documented exposures included: maternal diabetes (n=2 gestational, n=1 pre-gestational), viral infections (n=3, including 2 COVID-19), urinary tract infection requiring IV antibiotics (n=1), preeclampsia or pregnancy-induced hypertension (n=2), letrozole for infertility (n=1), selective serotonin reuptake inhibitor use (n=2), and cigarette smoking (n=3). No alcohol use was documented. Only one participant was conceived by assisted reproductive technology (in vitro fertilization). Data on preconceptual folate or prenatal vitamin use was incomplete, though many mothers reported taking prenatal vitamins at some point during pregnancy (Table 1). Due to limited documentation and small sample size, statistical analysis of exposure-RCEM relationships was not feasible.

### Genetic Testing

All participants underwent genetic evaluation and cytogenetic and/or molecular testing according to institutional protocols, availability, and clinical judgement, and testing therefore was not uniform across the cohort. About half (20/38) of the participants had a chromosomal microarray (CMA) (n=20), 17 had genome-wide sequencing (exome [ES] or genome sequencing [GS]), five had multigene panels (MGP), and three participants received single gene testing. The only diagnostic results were in the participants with monogenic conditions used as controls (R16, R17), which revealed their respective conditions (CFND and CODAS). All other testing was non-diagnostic, though variants of uncertain significance were reported in several cases (Table 1).

### Episignature Results

Figures 1–3 illustrate the clustering of the 40 new samples alongside the VACTERL/OAVS and control samples used for the EpiSign V5 RCEM episignature. Table 1 presents the methylation variant pathogenicity (MVP) scores for the 34 test samples. The majority of VACTERL samples, as well as participants R6, R7, R11, and R13 from the expanded RCEM cohort, showed methylation patterns consistent with the identified episignature. None of the participants with FND matched the VACTERL episignature.

**Figure 1:**
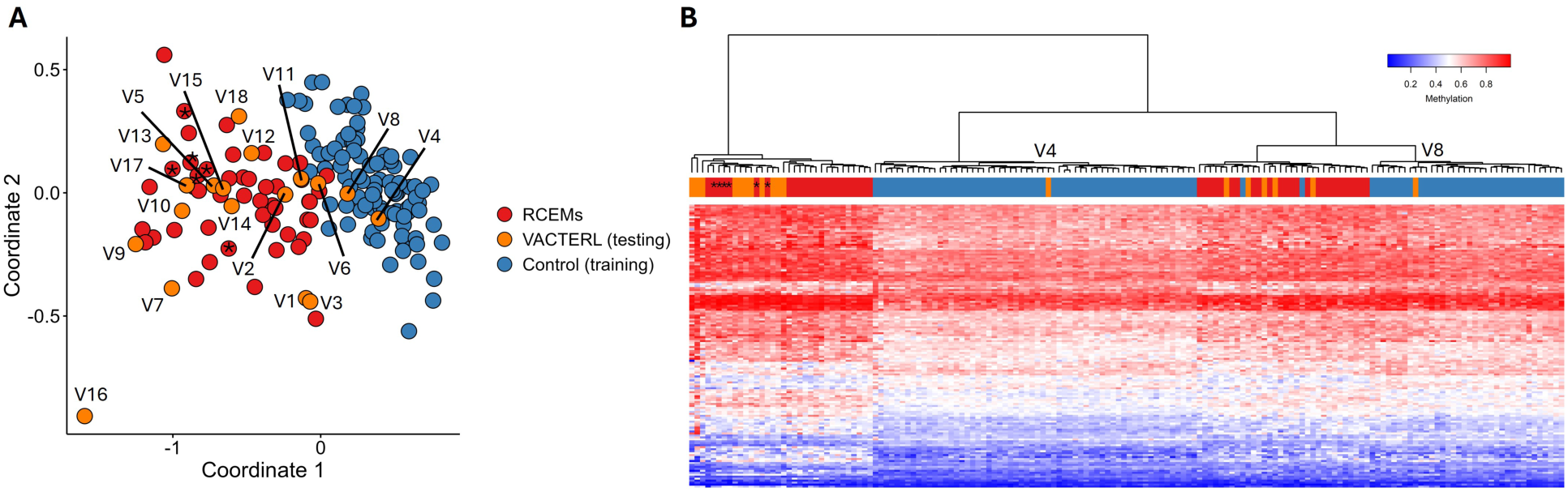
Multidimensional scaling (MDS) plots and heatmap for the VACTERL and partial VACTERL testing samples (orange) versus matched controls (blue). The EpiSign V5 RCEM episignature training cohort is included in red. The six VACTERL samples that were added to the training cohort are indicated with asterisks.

**Figure 2:**
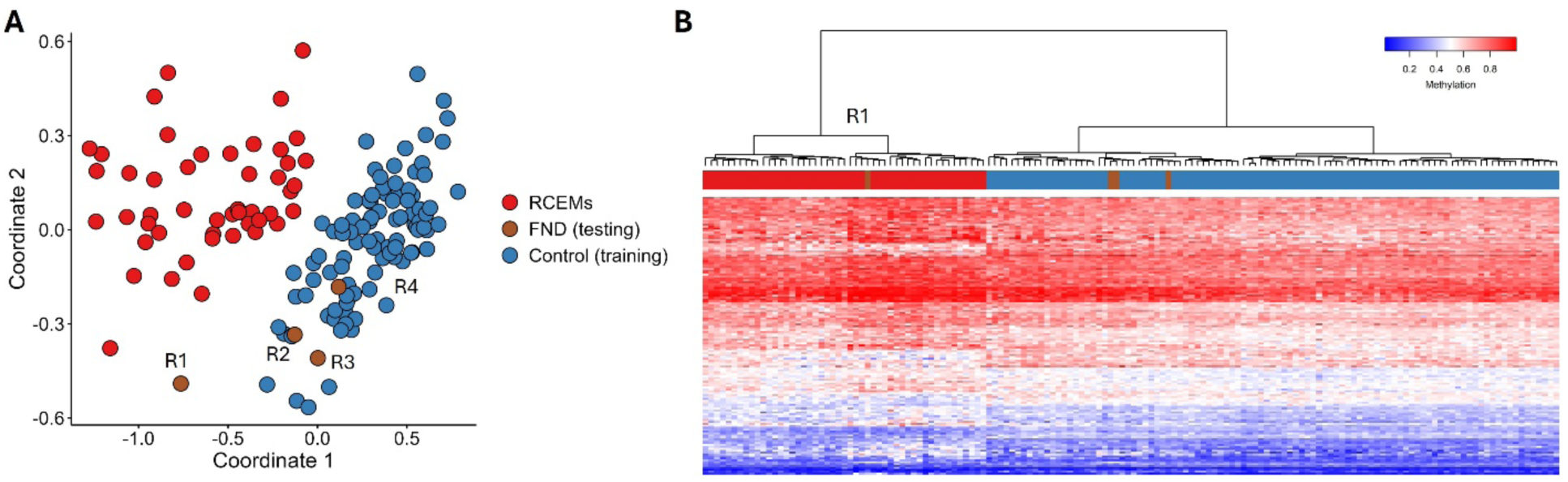
MDS and heatmap for the FND testing samples (brown) versus matched controls (blue) and the EpiSign V5 RCEM episignature (red).

**Figure 3:**
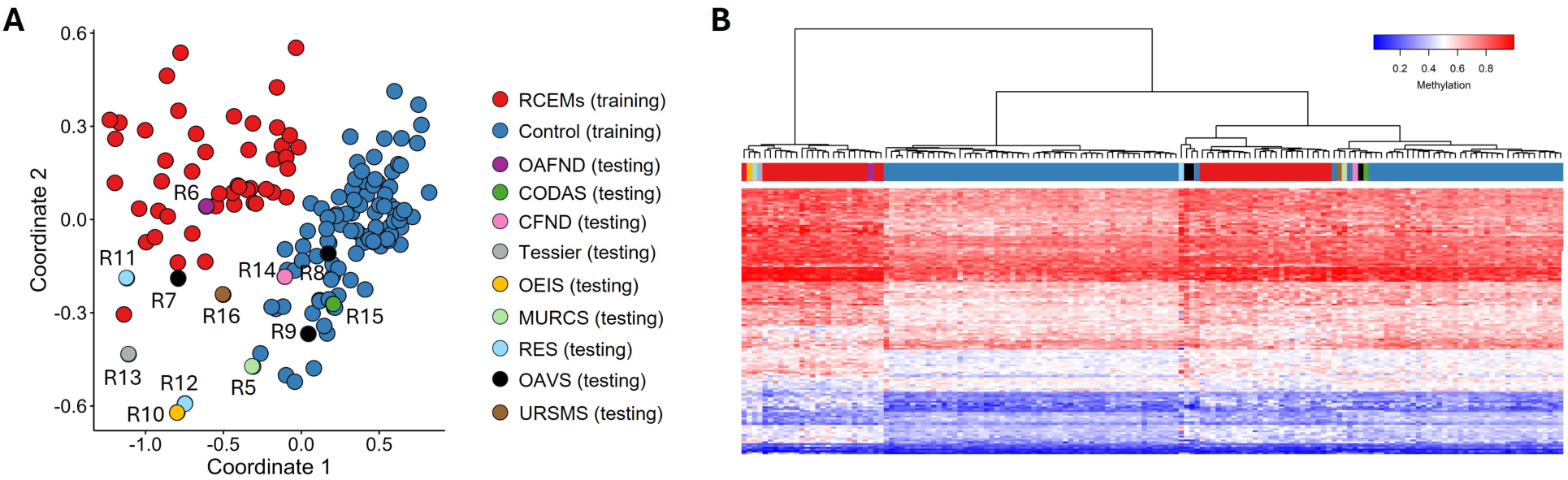
MDS and heatmap for the other RCEM-spectrum disorders (including OAFND, Tessier, OEIS, MURCS, RES, OAVS, and URSMS), along with the monogenic controls (CODAS and CFND), along with matched controls (blue) and the EpiSign V5 RCEM episignature (red).

Of the 21 samples from participants with VACTERL, all were positive for the previously established episignature: 18 robust (85.7%) and three intermediate (14.3%). Episignature sensitivity for participants with VACTERL (those with three or more features) was thus 100%. Six of these participants (V20-V25) showed such significant overlap with the previously demonstrated episignature that they were included as training cases (Figure 1, participants identified by an asterisk). Of the three participants with partial VACTERL presentations, two were negative (V4 and V8), and one (V3) had an intermediate episignature. One of these (V4) had a nondiagnostic GS.

The sample from the one participant with OAFND (R6) and one of two with RES (R11) showed robust episignature positivity, while the participant with Tessier cleft (R13) showed intermediate positivity (Figure 3). The second participant with RES (R12) did not cluster with the episignature, which is the child of the mother with VACTERL (V17, who did have the episignature), and neither did the four participants with FND (R1-4), nor the ones with MURCS (R5) and URSMS (R14). Of the three participants with OAVS, one showed intermediate positivity (nondiagnostic ES), while the other two were negative (one with non-diagnostic ES, the other with nondiagnostic CMA and MGP).

### Monogenic Controls

Two samples with known monogenic conditions were included as controls: CFND and CODAS syndrome. CFND shows significant phenotypic similarity to FND and OAFND, including hypertelorism, facial clefts, and craniosynostosis, but has a known molecular basis; pathogenic variants in *EFNB1*, following X-linked inheritance. CODAS syndrome is a rare autosomal recessive condition due to pathogenic variants in the *LONP1* gene with overlapping features including a median nasal groove (similar to FND/CFND). Notably, the features of the participant with CODAS initially resembled limb-body wall complex before exome sequencing revealed the molecular diagnosis. Both participants showed negative results for the EpiSign V5 RCEM episignature.

## DISCUSSION

In this independent and expanded cohort, we confirm and extend the findings of Haghshenas et al. (2024) by demonstrating that the established DNA methylation episignature is robust and reliable for VACTERL, while showing variable detection across other RCEM spectrum conditions. All individuals meeting clinical criteria for VACTERL were episignature-positive, with most showing robust positivity, whereas the two out of three participants with partial VACTERL/VACTERL-like presentations that did not meet full criteria had negative results. It is notable that the third participant with partial VACTERL with positivity for the intermediate episignature had tracheoesophageal fistula, which is considered to be a more specific feature of the association, while the other two had localized caudal defects. The participants with monogenic comparison conditions did not overlap with the established episignature, supporting specificity despite phenotypic resemblance.

This discovery of a stable, reproducible molecular biomarker in classically non-Mendelian, unsolved conditions raises fundamental questions about what these episignatures reflect, how they arise, and what their convergence across phenotypically distinct malformation syndromes reveals about shared disruptions in early embryonic development. DNA methylation episignatures differ fundamentally from sequence or structural variants in that they capture a downstream molecular state rather than primary genomic variation. In monogenic conditions, this state may reflect the functional consequence of a specific gene, domain, or pathway disruption. In RCEMs, the same concept may need to be broadened: an episignature may represent the stable molecular imprint of developmental disruption during a vulnerable embryonic window, produced by more than one upstream route. Its persistence in peripheral blood years after the malformations occurred speaks to the stability of selected methylation patterns and is central to their clinical utility (Kerkhof et al., 2024). This stability also raises important biological questions about when the signature is established, whether it contributes to pathogenesis or is a durable readout of earlier disruption, and why few individuals with overlapping clinical features or diagnoses do not carry the signature. We propose that the established episignature marks a period of shared epigenomic vulnerability during embryogenesis, arising from a combination of factors, rather than any single deterministic cause.

Prenatal exposure-associated episignatures provide a useful parallel. In fetal alcohol spectrum disorder, individuals meeting full diagnostic criteria demonstrated a distinct episignature, whereas those with partial or possible FASD showed partial overlap (Van der Laan et al, 2025). Alcohol exposure during pregnancy is hard to definitively confirm and may be continuous or limited to specific gestational periods. Timing is postulated to affect the development of clinical findings, such as typical facial features, and it appears that episignature development may correspond with presentation severity. Importantly, the absence of the episignature did not exclude the clinical diagnosis. Exposure to other teratogens, such as valproate, tends to be more discrete, well-documented and recognized. However, the fetal valproate episignature was not detected in every individual with confirmed exposure; 12 of 55 exposed individuals did not cluster with the episignature, and several of these had confirmed or suspected additional genetic etiologies (Haghshenas et al., 2024). These observations suggest that prenatal exposure is necessary but not always sufficient to generate a particular episignature and other modifiers may influence its emergence.

This imperfect concordance is also seen in episignature testing of mendelian conditions. Episignatures are generally highly specific, but sensitivity varies and is dependent on variant types and mechanisms represented in the episignature training cohort and the variants being tested. Individuals with pathogenic variants in a certain gene and negative results for its corresponding episignature may exist due to differing pathogenic mechanisms of those variants, and therefore a negative episignature result does not rule out a proposed clinical diagnosis or change the classification of a pathogenic variant (Kerkhof et al., 2024). Sensitivity has been listed as 40 to 100% across disorders (Husson et al., 2024), with a variable positivity rate by variant classification: 91% for likely pathogenic, 89% for pathogenic, and only 18% for variants of uncertain significance (Smits et al., 2025). The lower rate for VUSs likely reflects the known fact that only a subset of VUS’s will turn out to be truly causative with further functional or population data. This mirrors the pattern we observed in VACTERL, where episignature positivity is highest in clearly defined cases with specific features, and lower in partial or borderline presentations, where diagnosis is not clearly established.

Conversely, among episignature-positive individuals without a prior molecular diagnosis, subsequent exome or genome sequencing can identify causative variants in the proposed gene in a subset of cases (Kerkhof et al., 2024), 9/11 in Smits et al., (2025). These imperfect correspondences likely reflect a combination of platform sensitivity limits, threshold effects during critical developmental windows, or actual etiologic heterogeneity. Together, these observations show that episignature status is not a simple one-to-one surrogate for either genotype or exposure. Rather, it may mark a final common molecular consequence that can be reached through different mechanisms. For RCEMs, this is conceptually powerful: conditions long considered non-genetic or unsolved may not be etiologically uniform, but they may still share a measurable downstream biomarker.

OAVS is a particularly instructive case. Known historically under several names such as Goldenhar syndrome and hemifacial microsomia, OAVS is regarded as a heterogeneous group of craniofacial conditions unified by involvement of structures derived from the first and second branchial arches (Tingaud-Sequeira et al., 2022). Unlike most other RCEMs, OAVS is not uniformly non-genetic: some cases are attributable to defined chromosomal microdeletions, and monogenic subtypes have been identified (Tingaud-Sequeira et al., 2022). In this context, episignature-negative OAVS cases may represent true OAVS without the VACTERL/OAVS methylation pattern, phenocopies with alternative molecular etiologies, or subgroups in which timing, severity, or upstream mechanism differs. Expanded cohorts with systematic molecular investigation will be required to determine which fraction of clinically defined OAVS can be identified by the current episignature.

The variable episignature detection rates across the RCEM spectrum are also scientifically informative. RCEMs were grouped together based on overlapping clinical features across a spectrum, an absence of detectable genetic etiology, and a presumed early embryonic origin. (Adam et al., 2020; Mark, 2022). However, biological boundaries remain imprecise. Proposed mechanisms include NAD⁺ deficiency (Mark, 2022) and vascular disruption via vitelline steal (Stevenson, 2021). The identification of a DNA methylation episignature now offers an empirical molecular tool to test these boundaries: conditions that share the episignature may share a common pathogenic mechanism regardless of clinical label, while those that do not may be mechanistically distinct despite phenotypic overlap. In this light, the episignature positivity detected in OAFND and RES (conditions not originally classified as RCEMs) is striking. Whether these represent rare instances of a broader RCEM continuum or coincidental overlap will require validation in larger cohorts, but the finding suggests that the biological boundary of the RCEM epigenomic group may not coincide perfectly with its current clinical boundary. As episignature databases expand and cohorts grow, the methylation data themselves may help redefine the classification of embryonic malformation syndromes.

Beyond expanding RCEM cohorts, future investigations should systematically and prospectively capture prenatal exposure data. Our cohort demonstrates that such information is often incomplete in retrospective records, limiting our analysis. A small subset of congenital malformations already have established environmental determinants: periconceptual folate deficiency with neural tube defects, and maternal smoking with gastroschisis (Iskandar and Finnel, 2022, Baldacci et al., 2022). For RCEMs, systematic recording of maternal illness, medications, ART, placental complications, nutrition, infections, and other exposome variables would allow comparison of episignature-positive and episignature-negative individuals with the same clinical diagnosis. It would also help test whether certain exposures produce RCEM-like signatures only in the presence of permissive genotypes or other susceptibility states. The link with monozygotic twinning reinforces this concept: twinning itself, a mechanical, spontaneous event with no known genetic link, which changes the prenatal host environment, has a detectable DNA methylation episignature (van Dongen et al., 2021). RCEMs also show increased frequency and frequent discordance in monozygotic twins. This raises the possibility that twinning, placental environment, and methylation instability are not merely epidemiologic associations, but part of the same developmental susceptibility landscape.

## CONCLUSION

This study validates the recently established DNA methylation episignature as a robust biomarker for VACTERL, with all individuals with specific features testing positive, and those with negative results falling short of full diagnostic criteria. Reproducibility across independent cohorts and geographic regions supports its clinical utility for a diagnosis that has historically relied on exclusion and pattern recognition.

Variable sensitivity across the broader RCEM spectrum underscores that episignature testing must be interpreted in clinical context, with negative results not excluding a diagnosis, particularly for OAVS and less well-characterized conditions. Further exploration in LBWC, MURCS, OEIS complex, and pentalogy of Cantrell, as well as across an expanded list of similar conditions, will be necessary to determine the full extent of the shared epigenomic disruption, and may ultimately help redefine RCEM diagnostic boundaries on molecular rather than purely phenotypic grounds.

More broadly, these findings suggest that episignatures can reveal molecular convergence in conditions that are not explained by conventional genetic testing. While episignatures serve as functional adjuncts in Mendelian disease, they emerge as primary diagnostic anchors for RCEMs as diagnoses of exclusion. In RCEMs, a detectable methylation signature may represent the combined consequence of permissive genotypes, exposures, early embryonic events, or other modifiers acting within a vulnerable developmental window. This shifts RCEMs from purely descriptive diagnoses toward testable molecular entities and provides a platform for discovering the mechanisms that shape human malformation patterns.

## Data Availability

All data produced in the present study are available upon reasonable request to the authors

## Acknowledgements

We would like to thank all the participants and their families for their participation in our study.

This work was supported in part by Care4Rare SOLVE (Genome Canada OGI-147, CIHR GP1-155867, Ontario Research Fund). K.M.B. is supported by a CIHR Foundation Grant (FDN-154279) and a Tier 1 Canada Research Chair in Rare Disease Precision Health (CRC-2018-00175).

## Ethics Declaration

Participant data was obtained from multiple institutions, while the “main” Institutional Review Board (IRB) was housed within the corresponding authors’ institution: Western University Health Sciences Research Ethics Board, study number HSREB #106302. Additional approval was obtained from CHEO Research Ethics Board:11/04E and CTO-1577. All other institutions involved in this study received local IRB approval. Written informed consent was obtained from all individuals or guardians in accordance with protocols approved by the appropriate local human subject ethics committee at each academic/health sciences center.

## Artificial Intelligence (AI) Declaration

During the preparation of this work, Generative AI (Claude, Sonnet 5) was used for grammar, editing, and minimal re-wording/sentence restructuring in this document. All AI generated text has been carefully reviewed by the primary authors and co-authors, and the content was edited and reviewed as needed. The authors take full responsibility for the content of the publication.

